# The impact of COVID-19 disruption to cervical cancer screening in England on excess diagnoses

**DOI:** 10.1101/2020.11.30.20240754

**Authors:** Alejandra Castanon, Rebolj M Matejka, Francesca Pesola, Peter Sasieni

## Abstract

**Background:** Cervical cancer screening services in England have been disrupted by the COVID-19 pandemic.

**Methods:** Using routine statistics we estimate number of women affected by delays to screening. We used published research to estimate the proportion of screening age women with high-grade cervical intraepithelial neoplasia and progression rates to cancer. Under two scenarios we estimate the impact of COVID-19 on cervical cancer over one screening cycle (3y at ages 25-49 and 5y at ages 50-64). The duration of disruption in both scenarios is six months. In the first scenario all women have their screening interval is extended by six months. In the second some women (those who would have been screened during the disruption) miss one screening cycle, but most women have no delay.

**Results:** Both scenarios result in similar numbers of excess cervical cancers: 630 vs. 632 (both 4.3 per 100,000 women in the population). However the scenario in which some women miss one screening cycle creates inequalities - they would have much higher rates of excess cancer: 41.5 per 100,000 screened women compared to those with a six month delay (5.9 per 100,000 screened).

**Conclusion:** To ensure equity for those affected by COVID-19 related screening delays additional screening capacity will need to be paired with prioritising the screening of overdue women.

## Background

Cervical screening aims to identify abnormal cells in the cervix and treat them before they progress to cancer. In England, cervical screening is mostly carried out in general practice by specially trained nurses. Until December 2019, when the national roll out of HPV primary testing was completed, screening was done using liquid based cytology. Screening has been offered to women aged 25-49 at three-yearly intervals (although the plan is to extend to five-yearly with HPV testing) and to those aged 50-64 at five-yearly intervals. The age-appropriate coverage is around 72% of the eligible women.(1) In recent years, the European age-standardised incidence of cervical cancer in England for women aged 25-64 has been hovering around 9.5/100,000.(1)

Vaccination against HPV (bi-valent vaccine) was introduced in England in 2008–09 to girls aged 12–13 years (born from 1^st^ of September 1995, to 31^st^ of August 1996) and to a catch-up cohort aged 14–18 years (born from 1^st^ of September 1990, to 31^st^ of August 1995). Coverage among girls aged 12–13 years has remained around 86% since.(2)

Due to the COVID-19 pandemic cervical cancer screening has been severely disrupted the world over. In England, invitations for screening were suspended from April 2020.(3) Although call/recall was reinitiated in June, primary care providers were given the option to delay invitations for screening for up to six months if necessary.(4) Similar disruptions to screening have been reported in other countries.(5, 6) It is likely that the second and future waves of the pandemic will result in further disruption to screening.

As the pandemic is not yet over, it is currently unknown how quickly primary care and laboratories will be able to restore cervical screening services to pre-pandemic levels, what capacity will be available to address the screening backlog that has accumulated since GP practices closed for non-urgent face to face contact in March 2020 and how many women will be willing to take up their screening invitation in the post-COVID-19 era. Nevertheless, it is possible to estimate the size of the effect that a disruption like this is likely to have on cancer incidence, depending on the screening programme’s approach to compensate for the lost opportunity to be screened.

Assuming the health service will not be able to increase screening capacity considerably compared with previous years, it will be difficult or impossible to “catch-up” with the backlog of screening. Rather the service must choose between extending the screening interval for a whole round of the programme or trying to confine the disruption to women who have already missed being screened. Here we model the impact of delays to cervical cancer screening on excess diagnosis of cervical cancers among women of screening age (25-64 years) in England.

## Methods

The effect of a delay in attendance to cervical screening is explored under two scenarios. For both scenarios the impact of COVID-19 on the provision of cervical cancer screening would affect services over one screening cycle (3 years in women age 25-49 and 5 years in women aged 50-64) only. The length of the disruption is the same in both scenarios, but the proportion of women affected by the delay differs in each scenario. Both scenarios assume that follow-up of women testing positive at screening before the disruption were followed up on time and that follow-up services do not experience delays once the disruption ends.

The first scenario considers a rolling delay of six months for all women in England. This means that for a single screening cycle, women would be invited for HPV testing at 3.5 or 5.5 years after their previous invitation to cytology screening, depending on their age. They would resume with a standard interval thereafter. The age at which screening is offered would be permanently increased by 0.5 years (up to age 65.5y). Young women entering the programme after the disruption has been resolved would not be affected.

The second scenario assumes invitations to screening were likewise disrupted for a six-month period and that women who were due a screen during this period do not receive screening during this cycle (i.e. for 3 to 5 years depending on age). Women whose invitation was not affected by COVID-19 disruption would continue to be invited as normal.

Both scenarios assume no disruption to screening uptake once screening resumes.

### Population estimates

The age-specific numbers of women screened in England following an invitation for screening (in categories: “call” (first ever invitation to screening), “recall” (second and later routine invitations to screening) and “outside the programme” (predominantly women who were invited but attended 12 months or more after the invitation is issued)) as reported in the NHS Cervical Screening Programme statistics for year 2018-19(7) were used to estimate the numbers of women by age group who would have normally been routinely screened over a 12-month period. In the first scenario, the number of women screened in one year was multiplied by 3 or 5 depending on the age group to obtain the total population affected by the delay. In the second scenario the affected population was half of those attending over 12 months.

The national statistics was the source of data for the estimated size of the female population in mid-2019 in England by age group.(8)

### Population with cervical intraepithelial neoplasia

To estimate the number of screened women with a high-grade cervical intraepithelial neoplasia (CIN) detectable through HPV screening, data reported from the first round of screening in the English HPV primary screening pilot was used, where by far the majority of women were unvaccinated.(9) The pilot reported that 6.6% women at the age of 25-29, 1.6% at the age of 30-49 and 0.5% at the age of 50-64 years had a CIN grade 2 or worse detected following HPV primary screening, either at baseline or at one of two early recalls.

The proportion of women estimated to have high-grade CIN was multiplied by the total population affected by the disruption, in order to obtain the number of women in whom CIN2 or worse diagnoses were delayed because screening could not take place as scheduled. Estimates of the population affected were adjusted for the proportion of women aged 25-29 and 30-34 who were vaccinated with three doses. The proportions of the birth cohort who were vaccinated (and the age of vaccination) were taken from the national statistics(10) and are presented in supplementary table 1. The odds ratios of being diagnosed with a CIN grade 3 or worse by age at vaccination as reported for Scotland(11) (comparing to cohorts that were not offered vaccination) were used to adjust the proportion of high-grade CIN per 100,000 screened women to better reflect the detection in vaccinated cohorts. Compared to unvaccinated women, the odds of high-grade CIN were the lowest (OR=0.14) among those who were vaccinated at ages 12 or 13 years. The odds ratio increased thereafter to 0.18 in those vaccinated age 14 years, 0.29 at age 15 years, 0.27 at age 16 years, 0.55 at age 17 years and 0.85 at age 18 years.

### Progression of CIN

The proportion of high-grade CIN that would have progressed to cervical cancer in six months was estimated using progression estimates from the Landy et al.(12) modelling study. In that study, parameter sets were chosen to be consistent with the literature. They reported six-monthly progression rates from high-grade CIN to asymptomatic cancer of 0.12% for women aged <30 years, 0.25% at age 30-34, 0.35% at age 35-39, 0.65% at age 40-49, 0.9% at age 50-61 and 1.1% at age 62 years or more. For the second scenario, cumulative transition probabilities (following an exponential distribution)(13) up to 36 months (for women aged 25-49 years) or 60 months (for those aged 50-64) were calculated from the same six-monthly progression rates to estimate the cumulative number of women whose undetected CIN would have progressed to cervical cancer during one age-appropriate screening round (supplementary table 2).

## Results

Delaying screening for 6 months for the whole population (scenario 1) would result in approximately 10.7 million women being affected by the disruption, whereas delaying screening for an entire screening cycle (scenario 2) only to those directly affected would impact 1.5 million women (Table 1). However, both scenarios would result in very similar expected numbers of excess cancers, approximately 630 over one screening round. These cases amount to approximately 4 additional cases per 100,000 women in the general population over one screening cycle.

**Table 1.** Excess cancers due to delays in screening. Scenario comparison

|  | Scenario 1 | Scenario 2 |
| --- | --- | --- |
| Total population in England | 14,677,008 | 14,677,008 |
| Population affected by delayed screening | 10,699,491<sup>a</sup> | 1,522,219<sup>b</sup> |
| Excess cervical cancers | 630 | 632 |
| Rate of excess cervical cancer per 100,000 screened women | 5.9 | 41.5 |
| Rate of excess cervical cancer per 100,000 population | 4.3 | 4.3 |

<sup>a</sup> Women who undergo screening in one screening round of 3 or 5 years, depending on age, estimated from the NHS Cervical Screening Programme statistics for year 2018-19.
<sup>b</sup> Women who undergo screening during a period of 6 months, estimated from the NHS Cervical Screening Programme statistics for year 2018-19.

When only taking into account the women who were affected by the delay (i.e., those that would normally have participated in screening but were unable to do so, or could only do it with a delay), the excess cervical cancer incidence rates differed vastly between the two scenarios. Those who miss an entire round (scenario 2) would have seven times higher rates of excess cancer compared with those whose screening was delayed by 6 months (41.5 per 100,000 women compared to 5.9 respectively: Table 1).

The age-specific distribution of excess cancer diagnoses with a 6-month delay can be found in Table 2, and the results for 3- or 5-year delays are shown in Table 3. In both scenarios, women aged 40-49 years are expected to be the most affected whereas the impact to women aged 25-34 years is mitigated by vaccination against HPV 16 and 18.

**Table 2.** Scenario 1. Population affected and excess cancers given a rolling six-month delay to screening

| Age group in 2020 | Women affected by a 6-month rolling delay to screening<sup>a</sup> | Estimated number of women with CIN2/3 in population | Excess diagnoses of cervical cancer | Excess rates per 100,000 affected | Excess rates per 100,000 population |
| --- | --- | --- | --- | --- | --- |
| 25-29 | 1,533,957 | 45,772<sup>b</sup> | 55 | 3.6 | 2.9 |
| 30-34 | 1,329,927 | 18,952<sup>b</sup> | 47 | 3.6 | 2.5 |
| 35-39 | 1,365,366 | 21,846 | 76 | 5.6 | 4.1 |
| 40-44 | 1,238,154 | 19,810 | 129 | 10.4 | 7.5 |
| 45-49 | 1,316,637 | 21,066 | 137 | 10.4 | 7.3 |
| 50-54 | 1,698,585 | 8,493 | 76 | 4.5 | 3.9 |
| 55-59 | 1,284,705 | 6,424 | 58 | 4.5 | 3.1 |
| 60-64 | 932,160 | 4,661 | 51 | 5.5 | 3.2 |
| Total | 10,699,491 | 147,024 | 630 | 5.9 | 4.3 |

<sup>a</sup> Women who undergo screening in one screening round of 3 or 5 years, depending on age, estimated from the NHS Cervical Screening Programme statistics for year 2018-19.
<sup>b</sup>Rate of CIN2/3 in the population adjusted for proportion vaccinated.

**Table 3.**
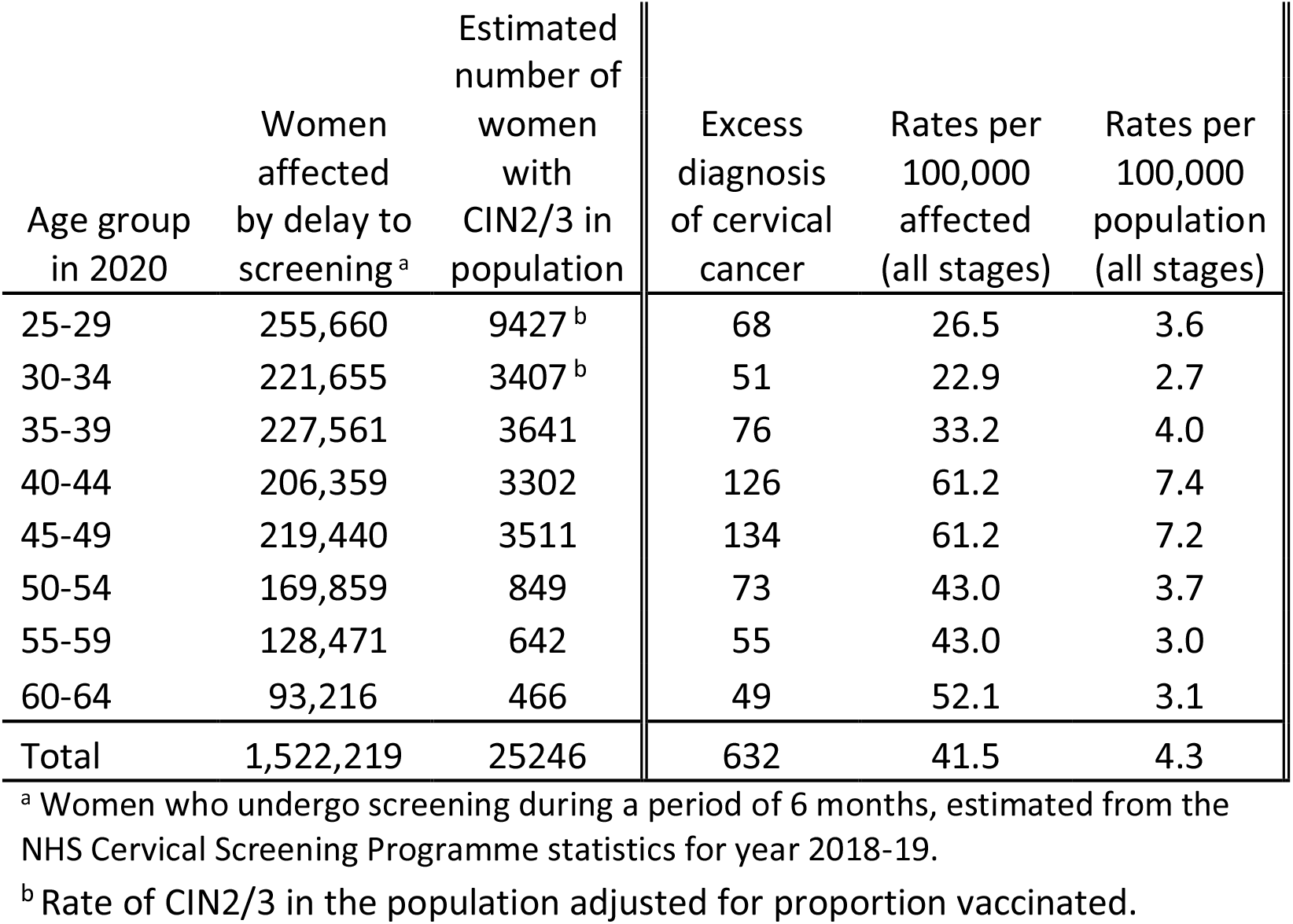
Scenario 2. Population affected and excess cancer due to women having their screen delayed by 3 or 5 years, depending on age.

| Age group in 2020 | Women affected by delay to screening <sup>a</sup> | Estimated number of women with CIN2/3 in population | Excess diagnosis of cervical cancer | Rates per 100,000 affected (all stages) | Rates per 100,000 population (all stages) |
| --- | --- | --- | --- | --- | --- |
| 25-29 | 255,660 | 9427 <sup>b</sup> | 68 | 26.5 | 3.6 |
| 30-34 | 221,655 | 3407 <sup>b</sup> | 51 | 22.9 | 2.7 |
| 35-39 | 227,561 | 3641 | 76 | 33.2 | 4.0 |
| 40-44 | 206,359 | 3302 | 126 | 61.2 | 7.4 |
| 45-49 | 219,440 | 3511 | 134 | 61.2 | 7.2 |
| 50-54 | 169,859 | 849 | 73 | 43.0 | 3.7 |
| 55-59 | 128,471 | 642 | 55 | 43.0 | 3.0 |
| 60-64 | 93,216 | 466 | 49 | 52.1 | 3.1 |
| Total | 1,522,219 | 25246 | 632 | 41.5 | 4.3 |

<sup>a</sup> Women who undergo screening during a period of 6 months, estimated from the NHS Cervical Screening Programme statistics for year 2018-19.
<sup>b</sup> Rate of CIN2/3 in the population adjusted for proportion vaccinated.

## Discussion

Cervical screening at regular intervals prevents cervical cancer.(14-16) The length of the interval typically represents the period during which the benefit of screening is observed.(15) Hence, any delays to cervical screening will negatively impact cancer diagnoses. Both scenarios modelled here resulted in about 630 excess cancers, equivalent to just over 4 per 100,000 women in the population, distributed across one round of screening (i.e. 3 or 5 years depending on age). However the risk of cervical cancer to an average woman who would have attended screening is seven times higher if they had to delay their screening for a whole screening round than if they had to delay screening for only 6 months.

It will be challenging to assess the impact of COVID-19 related delays by monitoring population rates of cervical cancer. The impact at a population level will be spread over a number of years. Further, given the roll-out of HPV primary screening in 2020 an initial increase in cancer diagnoses is expected (because the test is more sensitive than cytology).(17) Evaluation of screening histories from women diagnosed with cervical cancer once screening services resume will provide the best evidence of the actual impact of COVID-19 delays to cervical screening.

Our model can be applied to a variety of situations that have the ability to derail a cervical screening programme in the same way as has happened with the COVID-19 pandemic. A limitation of our analysis is that we had to rely on indirectly estimated parameter for the rate of progression of CIN to cervical cancer. This is not unusual in modelling studies, but the parameters used in this study had been previously calibrated to replicate cancer rates in England.(12) We have not taken into account the effect of any delays in diagnostics and treatment of women with a positive screen, nor any drop in the coverage once screening fully resumes; both would result in additional cases of cervical cancer. Finally, we focused on the additional burden of cancer due to the disruption. Any worsening of the prognosis of screen-detectable cancers due to a delayed diagnosis was beyond the scope of our analysis but would further worsen the impact of the COVID-19 disruption.

Our results show that the overall population burden of cervical cancer does not depend on whether scenario 1 or scenario 2 takes place. The two scenarios, however, clearly differ in terms of how this burden is distributed among the population. Scenario 2 culminates in a substantially higher excess risk per affected woman. Because the CIN2/3 lesions left undetected would have a longer time to progress (compared to scenario 1), it is also more likely that the excess cancers under this scenario would be diagnosed at later stages. In the name of equity, therefore, our analysis calls for measures that ensure that women do not miss an entire screening round on account of the COVID-19 disruption, i.e. scenario 2 should be avoided and scenario 1 would be preferable. Under naïve assumptions used in our model, this means that an additional 9 million women would be affected by the shorter disruption that is also not without a risk. However, this risk could be diminished in the programmes by (a temporary) increase in the screening capacity in order to clear the screening backlog (on top of the usual workload). If, for example, the screening capacity increased by 33%, it would still take 18 months to clear the 6-month backlog, but it would significantly decrease the total number of excess cancers. We note that a small increase in capacity is assumed in scenario 1 to ensure that women entering screening for the first time are not affected. Unfortunately, the demand for reagents to carryout HPV testing competes directly with the demands for COVID reagents and hence increasing screening capacity may not be feasible.

Increasing the screening capacity alone will not be sufficient. Making sure that women feel confident enough to attend for screening when they are due should be another priority. This is, again, highlighted by the seven-fold higher excess risk of cervical cancer among those who are unable or unwilling to access screening for a whole round, and highlights the importance of messaging to encourage women overdue their screen to attend as soon as possible. Unfortunately, it is often difficult for primary care providers to assess a woman’s prior screening history at the time when they are offered a screening appointment. Nevertheless consideration should still be given to strategies that will allow the identification and prioritization of screening of women affected by the COVID disruption to ensure that cervical screening remains a truly equitable service.(18)

## Data Availability

source data for this study is freely available in published literature.

## References

1. Screening & Immunisations team, NHS Digital. Cervical Screening Programme, England - 2018-2019. 2019 [Available from: https://digital.nhs.uk/data-and-information/publications/statistical/cervical-screening-annual/england---2018-19.

2. Public Health England (PHE). Human Papillomavirus (HPV) Vaccine Coverage in England, 2008/09 to 2013/14. A review of the full six years of the three-dose schedule. : Immunisation, Hepatitis and Blood Safety Department.; 2015[Available from: https://www.gov.uk/government/publications/human-papillomavirus-hpv-immunisation-programme-review-2008-to-2014.

3. Merrifield N. NHSE to start issuing cervical screening invitations again from this month. PULSE magazine, 3 June 2020;2020[Available from: http://www.pulsetoday.co.uk/clinical/clinical-specialties/cancer/nhse-to-start-issuing-cervical-screening-invitations-again-from-this-month/20040892.article.

4. Pearce C. NHS England is reviewing suspension of national screening programmes: PULSE magazine, 21 April 2020; 2020[Available from: http://www.pulsetoday.co.uk/clinical/clinical-specialties/cancer/nhs-england-is-reviewing-suspension-of-national-screening-programmes/20040657.article.

5. Khanna D, Khargekar NC, Khanna AK. Implementation of Early Detection Services for Cancer in India During COVID-19 Pandemic. Cancer Control. 2020;27(1):1073274820960471.

6. Cancino R, Su Z, Mesa R, Tomlinson G, Wang J. The Impact of COVID-19 on Cancer-Screening: Challenges and Opportunities. JMIR Cancer. 2020.

7. NHS Digtal. Cervical Screening Programme, England - 2018-19.; 2019.

8. Population estimates for the UK, England and Wales, Scotland and Northern Ireland: mid-2019 [Internet]. Office for National Statistics. 2020 [cited 02 September 2020]. Available from: https://www.ons.gov.uk/peoplepopulationandcommunity/populationandmigration/populationestimates/bulletins/annualmidyearpopulationestimates/mid2019estimates.

9. Rebolj M, Rimmer J, Denton K, Tidy J, Mathews C, Ellis K, et al. Primary cervical screening with high risk human papillomavirus testing: observational study. Bmj. 2019;364:240.

10. Annual HPV vaccine coverage in England in 2010/11 [Internet]. Department of Health. 2012 [cited 06 August 2019]. Available from: http://media.dh.gov.uk/network/211/files/2012/03/120319_HPV_UptakeReport2010-11-revised_acc.pdf.

11. Palmer T, Wallace L, Pollock KG, Cuschieri K, Robertson C, Kavanagh K, et al. Prevalence of cervical disease at age 20 after immunisation with bivalent HPV vaccine at age 12-13 in Scotland: retrospective population study. Bmj. 2019;365:1161.

12. Landy R, Windridge P, Gillman MS, Sasieni PD. What cervical screening is appropriate for women who have been vaccinated against high risk HPV? A simulation study. Int J Cancer. 2017.

13. Naber SK, Matthijsse SM, Rozemeijer K, Penning C, de Kok IM, van Ballegooijen M. Cervical Cancer Screening in Partly HPV Vaccinated Cohorts - A Cost-Effectiveness Analysis. PLoS One. 2016;11(1):e0145548.

14. Andrae B, Kemetli L, Sparen P, Silfverdal L, Strander B, Ryd W, et al. Screening-preventable cervical cancer risks: evidence from a nationwide audit in Sweden. Journal of the National Cancer Institute. 2008;100(9):622–9.

15. Sasieni P, Adams J, Cuzick J. Benefit of cervical screening at different ages: evidence from the UK audit of screening histories. British journal of cancer. 2003;89(1):88–93.

16. Lonnberg S, Anttila A, Luostarinen T, Nieminen P. Age-specific effectiveness of the Finnish cervical cancer screening programme. Cancer epidemiology, biomarkers & prevention : a publication of the American Association for Cancer Research, cosponsored by the American Society of Preventive Oncology. 2012;21(8):1354–61.

17. Dillner J, Rebolj M, Birembaut P, Petry KU, Szarewski A, Munk C, et al. Long term predictive values of cytology and human papillomavirus testing in cervical cancer screening: joint European cohort study. Bmj. 2008;337:a1754.

18. Sasieni P. Equality and equity in medical screening: what is fair? Lancet Gastroenterol Hepatol. 2019;4(8):578–80.

