## Supplemental Table 1 and 2 for "The impact of COVID-19 disruption to cervical cancer screening in England on excess diagnoses"

| Supplementary table 1. Proportion of the population protected against HPV 16/18 by age at vaccination and calendar year at fist screen. | | | | | | | | | | |
| --- | --- | --- | --- | --- | --- | --- | --- | --- | --- | --- |
| Vaccine age | Birth cohort | Coverage*  (three doses) | Protected against HPV 16/18 | Year enter screening | Proportion in the population aged 24-29yrs protected | | | Proportion in the population aged 30-34yrs protected | | |
|  |  |  |  |  | 2020 | 2021 | 2022 | 2020 | 2021 | 2022 |
| 12/13 | 01 Sept 95 – 31 Aug 97 | 80.9-84.4% | 100% | 2020-2022 | 0.17 | 0.34 | 0.51 | - | - | - |
| 14/15 | 01 Sept 94 – 31 Aug 95 | 75.7% | 95% | 2019 | 0.14 | 0.14 | 0.14 | - | - | - |
| 15/16 | 01 Sept 93 – 31 Aug 94 | 70.8% | 90% | 2018 | 0.13 | 0.13 | 0.13 | - | - | 0.13 |
| 16/17 | 01 Sept 92 – 31 Aug 93 | 48.1% | 85% | 2017 | 0.08 | 0.08 | - | - | 0.08 | 0.08 |
| 17/18 | 01 Sept 91 – 31 Aug 92 | 38.9% | 80% | 2016 | 0.06 | - | - | 0.06 | 0.06 | 0.06 |
| 17/18 | 01 Sept 90 – 31 Aug 91 | 47.4% | 80% | 2015 | - | - | - | 0.08 | 0.08 | 0.08 |

*Coverage obtained from 2010/11 vaccine coverage figures for England.(1)

| Supplementary table 2. Six monthly transition probabilities | | | | |  |  |
| --- | --- | --- | --- | --- | --- | --- |
| Months | Transition from high-grade CIN to stage IA preclinical cervical cancer | | | | | |
|  | <30 | 30-34 | 35-39 | 40-49 | 50-59 | 60+ |
| 6 | 0.12% | 0.25% | 0.35% | 0.65% | 0.90% | 1.10% |
| 12 | 0.24% | 0.50% | 0.70% | 1.29% | 1.78% | 2.18% |
| 18 | 0.36% | 0.75% | 1.04% | 1.93% | 2.66% | 3.25% |
| 24 | 0.48% | 1.00% | 1.39% | 2.57% | 3.54% | 4.30% |
| 30 | 0.60% | 1.24% | 1.73% | 3.20% | 4.40% | 5.35% |
| 36 | 0.72% | 1.49% | 2.08% | 3.82% | 5.26% | 6.39% |
| 42 | 0.84% | 1.73% | 2.42% | 4.45% | 6.11% | 7.41% |
| 48 | 0.96% | 1.98% | 2.76% | 5.07% | 6.95% | 8.42% |
| 54 | 1.07% | 2.22% | 3.10% | 5.68% | 7.78% | 9.43% |
| 60 | 1.19% | 2.47% | 3.44% | 6.29% | 8.61% | 10.42% |

1.	Annual HPV vaccine coverage in England in 2010/11 [Internet]. Department of Health. 2012 [cited 06 August 2019]. Available from: <http://media.dh.gov.uk/network/211/files/2012/03/120319_HPV_UptakeReport2010-11-revised_acc.pdf>.
